# Cell-type specific transcriptional risk scores and longitudinal multiple sclerosis outcomes

**DOI:** 10.64898/2026.09.21.26363545

**Authors:** Mae Upcott, Beili Shao, Nicholas Bray, Sam Loveless, Emma C. Tallantyre, Neil P. Robertson, Karim L. Kreft

## Abstract

Transcriptional risk scores, an integrated score of genomic susceptibility variants with expression Quantitative Trait Loci (eQTL) data, may more accurately distinguish clinical phenotypes compared to polygenic risk scores. This approach has not been applied to multiple sclerosis. We integrated data from the largest GWAS of MS, involving 14,802 MS patients, with eQTL data from 7,466 blood donors and 5,494 brain donors, using summary-data-based Mendelian randomization (SMR) followed by the heterogeneity in dependent instruments (HEIDI) test. MS SNVs with a significant eQTL were integrated with single-cell RNA sequencing data from 1,27 million peripheral blood mononuclear cells from 927 donors and 750,614 single nucleus RNA sequencing central nervous system cells from 192 individuals using Bayesian colocalization. Derived polygenic and cell-line specific transcriptional risk scores were compared with age-related MS severity score (ARMSS) in a cohort of 1,077 MS patients from the South Wales MS Registry and with MRI-derived age-adjusted total brain volumes of 272 MS patients within the UK Biobank. Cell-line specific transcriptional risk scores were associated with MS outcomes in two independent cohorts. Our results identified MS-associated eGenes enriched across several biological pathways, providing insights into pathophysiological mechanisms in MS that could facilitate precision medicine and identify novel druggable pathways.

## Introduction

Predicting disease severity in multiple sclerosis (MS) at disease onset remains elusive, due to the subsequent disease heterogeneity. Estimates suggest that single nucleotide variants (SNVs) explain approximately 19% of the heritable component of MS susceptibility (International Multiple Sclerosis Genetics Consortium, 2019) and 13% of disease severity (International Multiple Sclerosis Genetics Consortium and MultipleMS Consortium, 2023). Previous studies have shown that MS susceptibility SNVs are mainly immune related (International Multiple Sclerosis Genetics Consortium, 2019), whereas SNVs associated with MS severity are predominantly expressed in the central nervous system (International Multiple Sclerosis Genetics Consortium and MultipleMS Consortium, 2023; Jokubaitis et al., 2023).

Polygenic risk scores (PRS) summate the effect of multiple SNVs to estimate an individual’s total genetic predisposition to a trait, but currently PRS lack the ability to sensitively distinguish clinical groups (Shams et al., 2023). Moreover, utilisation of single disease-specific SNVs from MS genome wide association studies (GWAS) to predict long-term severity has proven to be challenging (Kreft et al., 2024).

In Alzheimer’s and Crohn’s disease, integrating GWAS with gene expression data (eQTL) to derive a transcriptional risk score (TRS) has improved the sensitivity to detect clinically relevant subgroups of patients compared to traditional PRS (Marigorta et al., 2017; Pyun et al., 2024). This approach has not previously been applied to MS cohorts.

Therefore, we leveraged summary level MS susceptibility GWAS data with eQTL data and single nuclei RNA sequencing data to produce TRS, reflecting gene expression regulated by SNVs in a cell-type specific manner. These scores were correlated with important disease outcomes in a deeply-phenotyped cohort of disease modifying treatment (DMT) naive MS patients (n=1,077) of the South Wales MS registry and in an independent cohort with brain volume measurements as a proxy of atrophy and MS severity in the UK Biobank (n=272).

## Materials and methods

### SNV identification for transcriptional risk scores using integration of GWAS with eQTL data

Summary data-based Mendelian Randomisation (SMR) allows integration of GWAS data with eQTL data to prioritise putative causal genes (Zhu et al., 2016). We integrated the summary statistics of the discovery phase of the largest MS susceptibility GWAS consisting of 14,802 MS patients and 26,703 unaffected individuals (International Multiple Sclerosis Genetics Consortium, 2019) with publicly available eQTL data from a total of 7,466 blood donors (GTEx Consortium. 2020; Lloyd-Jones., 2017; Lappalainen et al., 2013; Westra et al., 2013) and 5,494 brain donors (Lappalainen et al., 2013; Qi et al 2018; Qi et al., 2022; Wang et al., 2018., Yazar et al., 2022). We applied default thresholding for SMR-analysis as previously described (Zhu et al., 2016). In brief, eQTLs with a p-value <5*10^-8^ were selected with a R^2^ between 0.05 and 0.90 with the top GWAS SNP at each locus. SMR analysis was used to identify gene expression differences associated with MS risk alleles at a false discovery rate (FDR)-corrected p-value of <0.05. The Heterogeneity In Dependent Instruments (HEIDI) test was subsequently used to distinguish those associations that are likely to arise from linkage disequilibrium, with those with HEIDI p-values ≤0.1 excluded from further analysis.

### Bayesian colocalization

We applied Bayesian colocalization to significant susceptibility SNVs identified by SMR/HEIDI, leveraging publicly available single-cell RNA sequencing (scRNA-seq) data. The peripheral blood mononuclear cell (PBMC) dataset comprised 1.27 million cells from 927 donors, including B lymphocytes, CD4+ and CD8+ T cells, dendritic cells, monocytes, natural killer cells, and plasma cells (Yazar et al., 2022). The central nervous system (CNS) dataset comprised single-nucleus RNA sequencing (snRNA-seq) data from 750,614 cells across 192 individuals, including astrocytes, endothelial cells, excitatory neurons, inhibitory neurons, microglia, oligodendrocytes (ODCs), oligodendrocyte precursor cells (OPCs), and pericytes (Bryois et al., 2022). Posterior probability (PP.H4) >0.80 was considered as evidence that a SNV is associated with an eQTL (eGenes). Identified eGenes were classified as either immune, CNS or both depending on the cell or tissue type associated with the eGene. We included cell-line specific TRS for immune and CNS cells.

### Genotyping and extracting target SNVs from South Wales MS Registry cohort

Welsh cohort genotyping methodology has previously been described (Kreft et al., 2024). In summary, DNA was extracted from blood samples collected in EDTA-containing tubes (BD) and stored at -80°C. Genotyping was conducted using either the Illumina Infinium CoreExome-24 v2 or v3 arrays following the manufacturers’ protocols. Stringent quality control measures were applied, and genotype data were imputed as previously described (Kreft et al., 2024). Genome-wide significant susceptibility SNVs identified by SMR/HEIDI analysis were extracted. If an eGene SNV could not be identified within in the South Wales MS Registry genotype data, we assessed whether we could use a proxy SNV with an R^2^=1 in the CEU population using publicly available data (https://ldlink.nih.gov/). We have used five proxy SNVs in perfect linkage disequilibrium, but we were unable to find a proxy SNV for 47 out of 387 of SNVs identified using SMR/HEIDI (See Supplementary Table S1& S2).

### Calculation of polygenic and transcriptional risk scores

To compute PRS, we applied SBayesRC, a Bayesian approach leveraging genomics with functional annotations (Zheng et al., 2024) and calculated separate polygenic risk scores using previously published GWAS of MS Susceptibility and Progression (International Multiple Sclerosis Genetics Consortium, 2019, International Multiple Sclerosis Genetics Consortium., 2023).

For the calculation of TRS, beta coefficients of each eQTL association were standardised to a mean of 0 and a standard deviation of 1. The MS susceptibility or progression risk allele of a SNVs was used to determine the direction of risk. We subsequently polarised transcriptomic values in the direction of the MS genomic risk allele, maintaining alignment to previous studies (Marigorta et al., 2017; Pyun et al., 2024). TRS were calculated by summating all polarized beta-coefficients across identified variants. All PRS and TRS were subsequently transformed into z-score to allow comparison across scores. QQ-plots were generated to visually assess if the scores followed a Gaussian distribution (See Supplementary Fig. S1).

### UK Biobank data extraction

Imputed genetic, imaging, and phenotypic data were accessed under an approved UK Biobank application (ID 761882) and analysed within the UK Biobank Research Analysis Platform. Whole-brain tissue volume was derived from the pre-existing UK Biobank structural MRI pipeline (Data-Field 25009, grey and white matter combined, normalised for head size) from the first imaging visit. MS cases were defined using the ICD-10 diagnostic code G35. TRS were calculated similarly to the South Wales MS dataset. Associations between TRS and brain volume were evaluated using linear regression models adjusted for age.

### Ethics

The Research Ethics Committee (REC) of Health and Care Research Wales (05/WSE/03/111,19/WA/0289 and 24/WA/0049) and the North West Multi-centre REC for UK Biobank (16/NW/0274) approved the studies, and all participants provided written informed consent before inclusion.

### Gene-set enrichment analysis

We performed pathway analysis on eGenes stratified according to immune and CNS function using GSEA analysis (Subramanian et al., 2005). FDR adjusted p-values

<0.05 were considered as evidence of overrepresentation.

### Gene expression in CSF and post-mortem tissues

We determined single cell gene expression of SMR/HEIDI identified genes in non-inflammatory tissues (Emani et al., 2024). Moreover, we determined for each significant eGene whether this gene is differentially expressed in CSF of MS patients versus controls (Jacobs et al., 2025) and in post-mortem MS tissue versus non-MS controls (MacNair et al., 2025). We considered DEG statistically significant with an FDR p<0.05 in the respective studies.

### Clinical outcomes and statistical analysis

To prevent confounding-by-indication (patients with unfavourable prognostic factors are more likely to have worse outcomes and to receive high-efficacy disease modifying treatments, which may bias long-term disease outcomes), we included only a DMT naive subgroup of the South Wales MS Registry (n=1,077). Cohort demographic and clinical characteristics were provided using descriptive statistics.

Last-recorded Age-related Multiple Sclerosis Severity (ARMSS) was calculated and stratified into quartiles and Kruskal-Wallis test with FDR adjustment was used to compare risk scores between ARMSS quartiles. Associations between the risk scores and the rank inverse nominal transformation allowing a Gaussian distribution of last-recorded ARMSS were assessed using a multivariate linear regression model adjusted for sex and the number of relapses during the first five years of disease. Time between 1^st^ and 2^nd^ relapse was stratified into less or greater than 2 years and t-tests with FDR adjustments were used to compare risk scores. This timeframe was selected as relapse frequency in the 2-year period after disease onset has been associated with greater disease severity (Scalfari et al., 2010).

For the analysis of age at disease onset, all pwMS with onset less than 19 years old were excluded due to limited number of patients. Age of disease onset was stratified into 10-year epochs and Kruskal-Wallis test with FDR correction between age strata. Statistical significance was determined as FDR-adjusted p-value <0.05. A flowchart of the study is provided in figure 1.

**Fig. 1.**
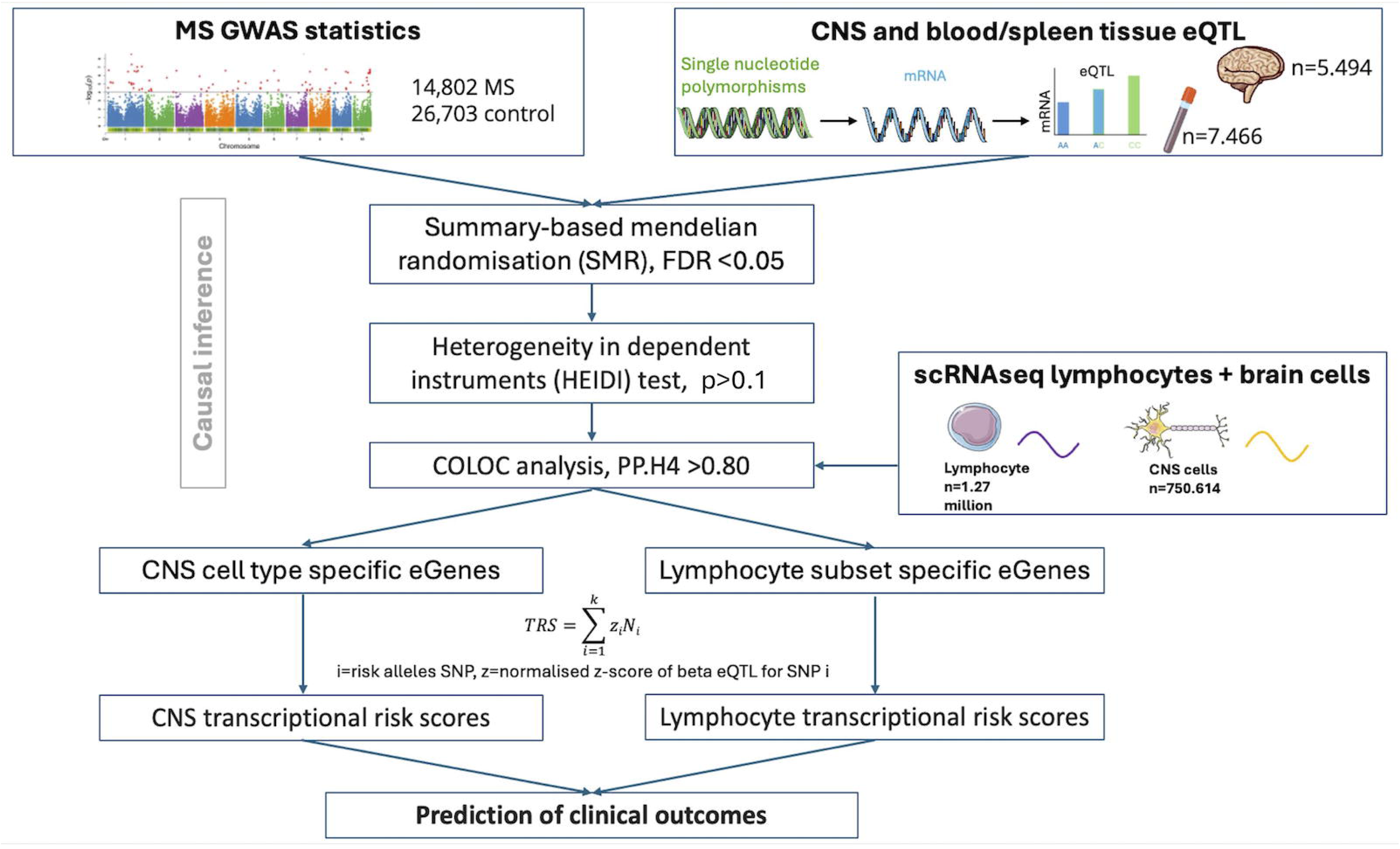
Flowchart of study.

All analyses were done with R version 4.2.1 (in Rstudio 2022-06-23 ucrt), R packages included tidyverse, corrplot, ggsignf, ggplot2, dplyr and lubridate, coloc (5.3.2), SMR (1.3.1).

## Results

### We identified 88 CNS specific and 109 immune specific genes associated with MS susceptibility

In total, we identified altered gene expression of 240 significant (eGenes) after SMR and HEIDI filtering associated with MS susceptibility, of which 43 were overlapping between immune and CNS tissues. Interestingly, 77 of the 88 CNS specific genes were not previously associated with MS, and 85 of 109 immune genes are newly identified (See Supplementary Table S3 & S4 respectively) We then performed gene set enrichment analysis. CNS genes were linked to kinase activity and immune signalling, potentially indicating the importance of neuro-immune interactions (Fig. 2A), whereas immune genes were associated with lymphocyte biology (Fig. 2B).

**Fig. 2.**
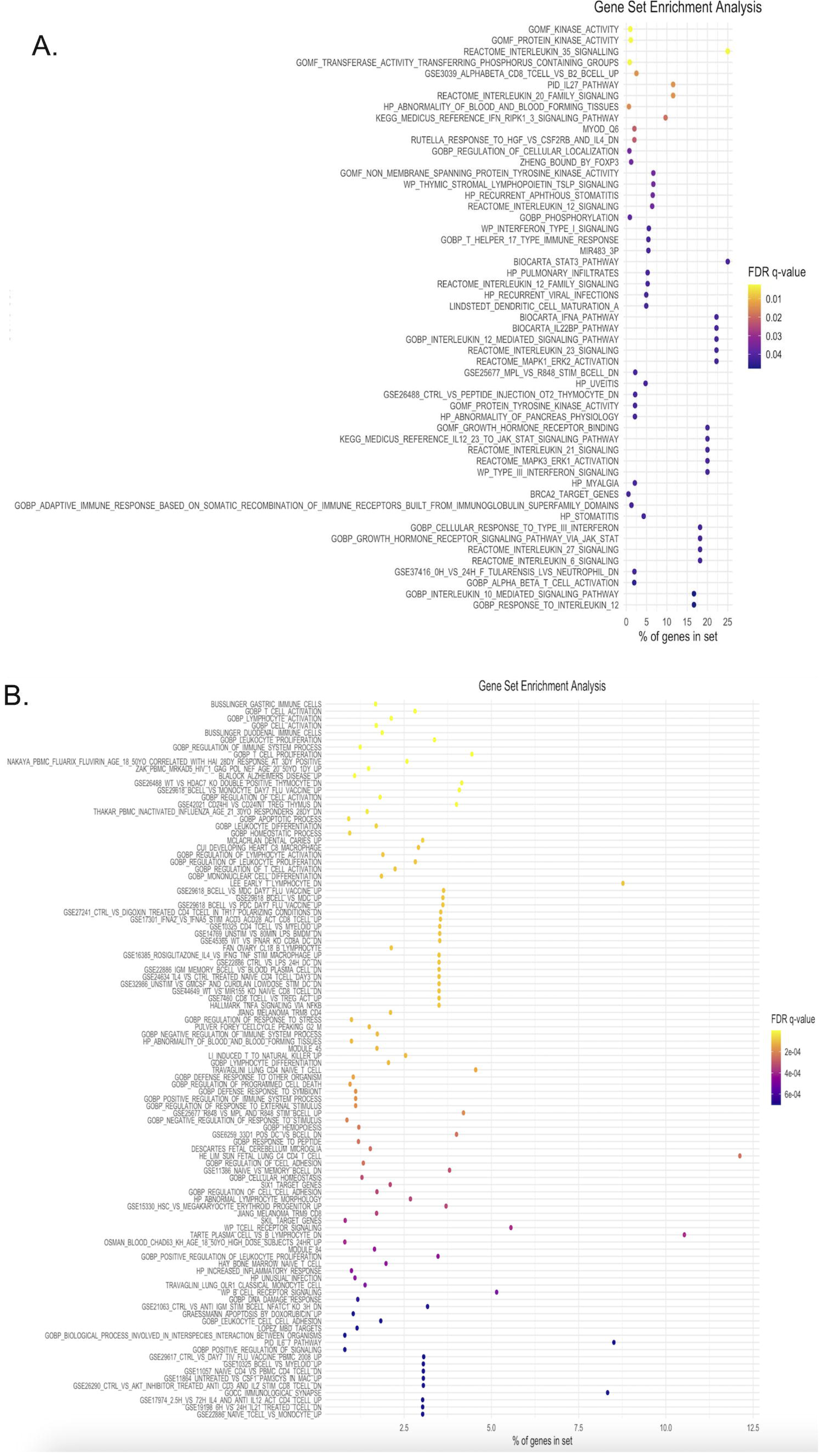
Gene Set Enrichment Analysis (GSEA) on significant eGenes after SMR and HEIDI filtering associated with MS susceptibility. Gene Set Enrichment Dot plots showing biological pathways and gene sets enriched in **A:** CNS and **B:** Immune Specific MS Susceptibility e-genes.

### MS-related genes are differentially expressed in CSF and post-mortem lesions

To further confirm the relevance of these newly identified genes in the CNS, we assessed if the eGenes are expressed in a cell-type specific manner in non-inflammatory post-mortem brain tissue (Emani et al., 2024). Interestingly, we observed the highest percentage of cells expressing of these genes in inhibitory and excitatory neurons (Fig. 3A). Next, we determined if those genes are differentially expressed in MS post-mortem lesions (Macnair et al., 2025). The majority of the identified CNS genes are indeed differentially expressed in both white and grey matter lesions in a cell-type specific manner in MS (Fig. 3B/C). Finally, we assessed if the newly identified immune genes are differentially expressed in MS peripheral blood and CSF immune cells (Jacobs et al., 2025). A high number of identified genes are in a cell-type specific manner differentially expressed in either blood or CSF. Some genes are differentially expressed in several immune cell subsets (Fig. 3D). Taken together, the newly identified genes using SMR are plausible candidates to be involved in MS pathogenesis.

**Fig. 3.**
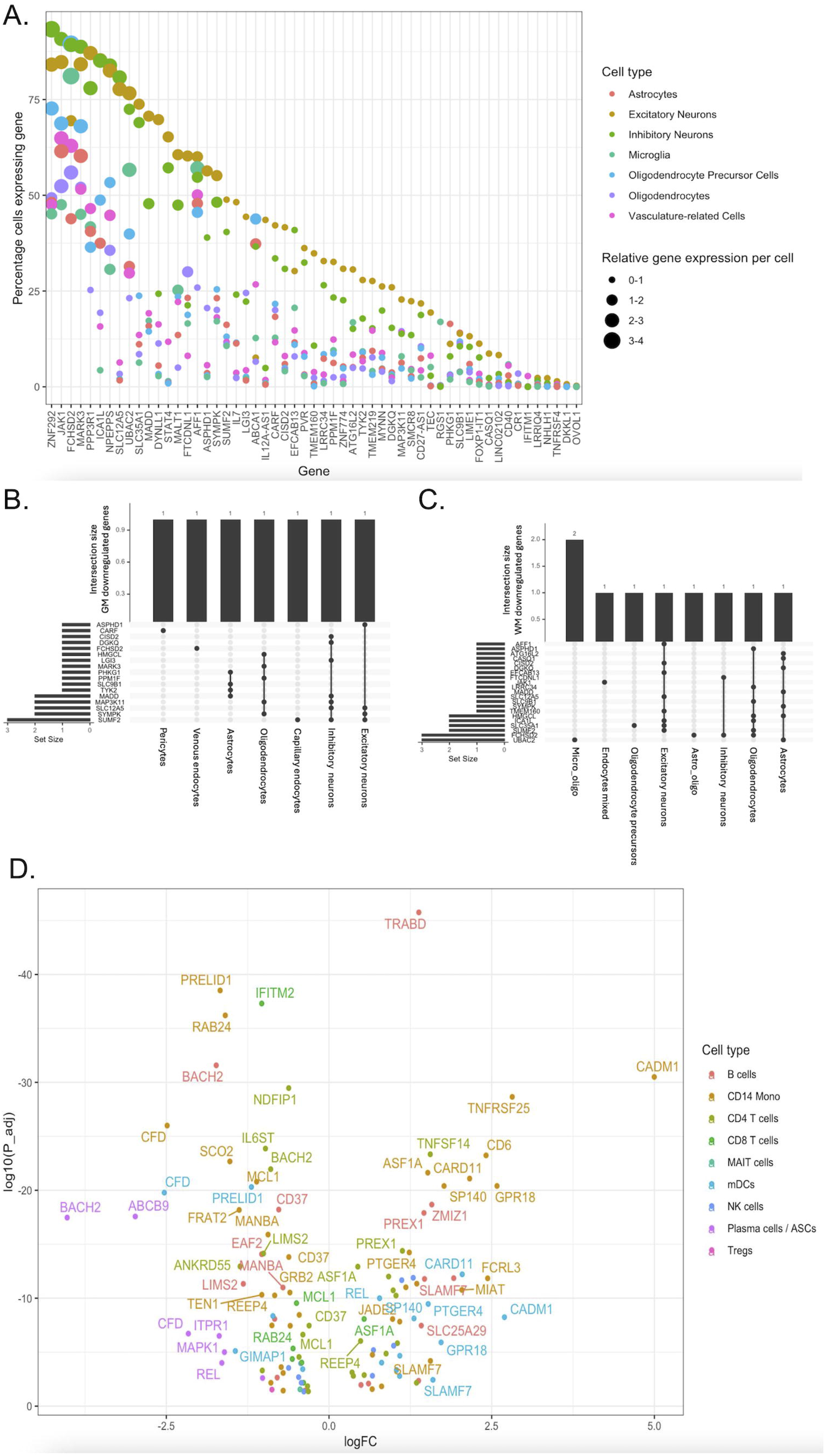
Cell-type–resolved gene expression and intersection analyses across immune cell populations. **A:** Dot plot showing relative expression of genes across CNS cell types in non-inflammatory control tissue. **B-C:** UpSet plots summarizing differentially expressed genes across CNS cell types in MS lesions. **D:** Volcano plot of differentially expressed genes across immune cell types comparing blood versus CSF of MS patients.

### Welsh longitudinal prospective MS cohort

Next, we compared the risk scores between clinical subgroups. We included 1,077 treatment naive MS patients, of which 747 (69.4%) were female with a median age of onset of 32.2 (IQR 16) years. 143 (13.3%) of people living with MS (pwMS) had a diagnosis of primary progressive MS. 512/633 (80.9%) of the participants who underwent CSF analysis were oligoclonal band positive. The median last recorded ARMSS score for the cohort was 6.2 (IQR 4.6, Table 1).

**Table 1:** Demographics and clinical characteristics.

|  |  |
| --- | --- |
| Total cohort | 1,077 |
| Sex F (n, %) | 747 (69.4%) |
| Median age at onset (IQR) | 32.2 (IQR 16) |
| OCB +ve | 512/633 (80.9%) |
| PPMS | 143 (13.3%) |
| Median last recorded ARMSS (IQR ) | 6.2 (IQR 4.6). |
| Median follow-up duration (years) (IQR) | 19.8 (18) |

OCB: oligoclonal bands only present in cerebrospinal fluid

### Transcriptional, but not polygenic risk scores are associated with MS outcomes

In patients with relapsing-onset MS, total and immune tissue TRS were associated with time between 1^st^ and 2^nd^ relapse (FDR adjusted p=0.02 and 0.02). This association was not found with PRS (Fig. 4). Both total, immune and CNS tissue TRS were associated with age of disease onset (FDR adjusted p=0.006, 0.05 and 0.02 resp.) and susceptibility PRS were also associated with age of disease onset (FDR adjusted p=0.04, Fig. S4A-E).

**Fig. 4.**
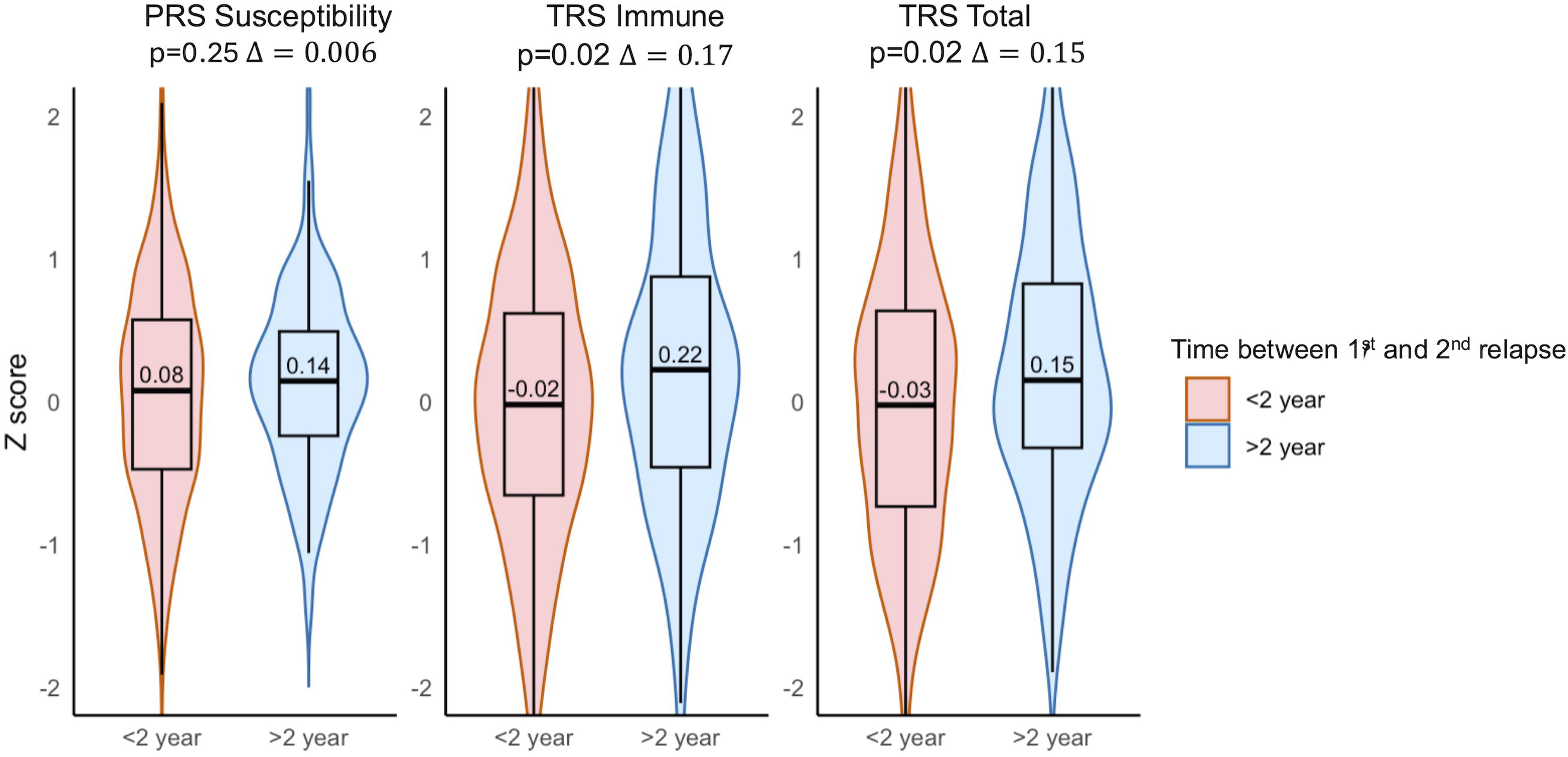
Violin plots of polygenic susceptibility risk scores, immune and CNS transcriptional risk scores, and time between first and second relapse, stratified by ≤2 years versus >2 years.

We did not observe an association between ARMSS and MS susceptibility PRS (FDR p=0.46) or progression PRS (FDR p=0.35, Fig. S3A-B.). Total TRS were also not associated with last recorded ARMSS (FDR p=0.07, Fig. S3C). However, the immune TRS was associated with last recorded ARMSS (FDR p=0.025, Fig. 5A), while we did not observe an association with CNS TRS (FDR p=0.19, Fig. S3D). Patients in the highest quartile of last recorded ARMSS had a significantly greater immune TRS compared to those in the lower quartile (Post hoc Dunn test (p= 0.0015). Linear regression adjusted for number of relapses in the first 5 years and sex as potential confounders confirmed that immune TRS were correlated with last-recorded ARMSS (p=0.039, t=2.06). A one unit increase in the immune transcriptional risk Z-score is associated with a 7% increase in ARMSS score (β regression coefficient=0.07, 95% CI 0.003-0.13, Fig 5B).

**Fig. 5.**
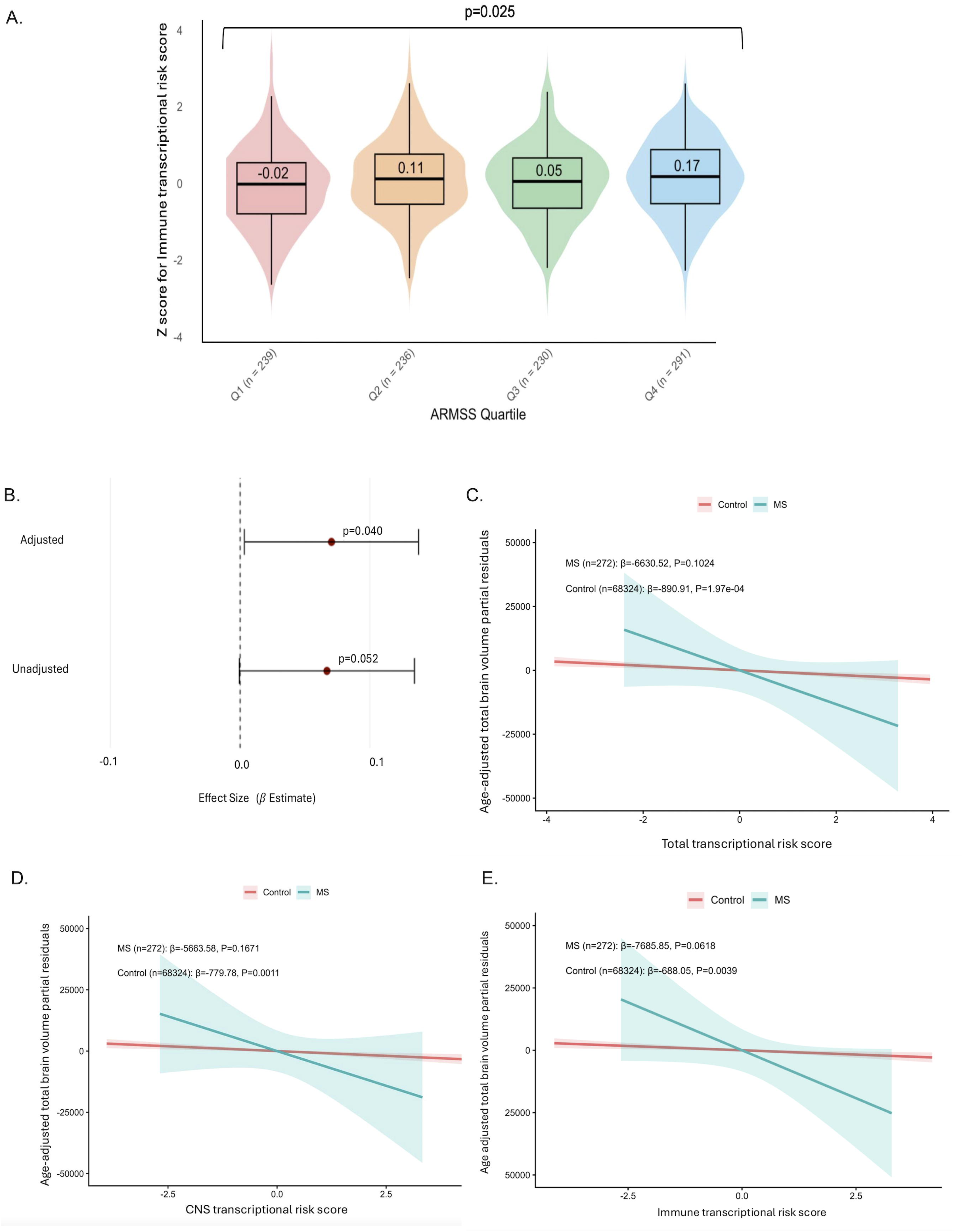
**A-B:** Immune transcriptional risk score and last recorded ARMSS A: Violin plot of Immune transcriptional Risk Score and last recorded ARMSS stratified into quartiles. **B:** Forest plot showing effect size estimates from linear regression of the Immune transcriptional Risk Score on last recorded ARMSS (unadjusted and adjusted for number of relapses in the first 5 years and sex) **C-E:** Linear regression of standardised transcriptional risk scores and age-adjusted total brain volume (TBV) in UK Biobank participants, stratified by multiple sclerosis (MS) and controls **C:** Total transcriptional risk score and total brain volume. **D:** CNS transcriptional risk score and total brain volume. **E:** Immune transcriptional risk score and total brain volume

Next, we wanted to validate the association between MS Severity (ARMSS) and TRS. Although the UK Biobank does not hold granular details on MS disability trajectories, it has detailed MR imaging data on total brain volumes. Brain atrophy in MS is a well-validated proxy for MS severity (Matthew et al,. 2023). We observed a significant inverse correlation between total, CNS and TRS and total brain volume of 68,324 participants from the general population (beta -890.91, p=1.97*10^-4^, beta - 779.78. p=0.0011, beta -688.05, p=0.0039). In the 272 pwMS in the UK Biobank, we observed a directionality concordant result with larger effects sizes, although the results were borderline non-significant, most likely due to relatively limited power (Fig. 5C-E). Nevertheless, the results indicate that higher TRS are associated with smaller brain volumes in the general population, with a more pronounced effect in MS, likely associated with a more severe disease.

### Cell-line specific risk scores and MS disease severity (ARMSS)

Next, we computed cell-type specific TRS for both central nervous system and immune cell types. B-lymphocyte and CD8+ cytotoxic T-lymphocyte TRS were only associated with last recorded ARMSS (resp. p=0.037 and 0.005, Fig. 6A-B). Of note, B lymphocyte and CD8+ T-lymphocyte TRS were highly correlated (Spearman’s rho=0.87). In the UK Biobank, CD8+ T-lymphocyte, but not B-lymphocyte (Fig. 6C) TRS were associated with age-adjusted total brain volume in controls with a non-significant but directionality concordant results in pwMS (beta -718.61, p=0.002, Fig. 6D), further underscoring the importance of this score in long-term outcomes of MS severity.

**Fig. 6.**
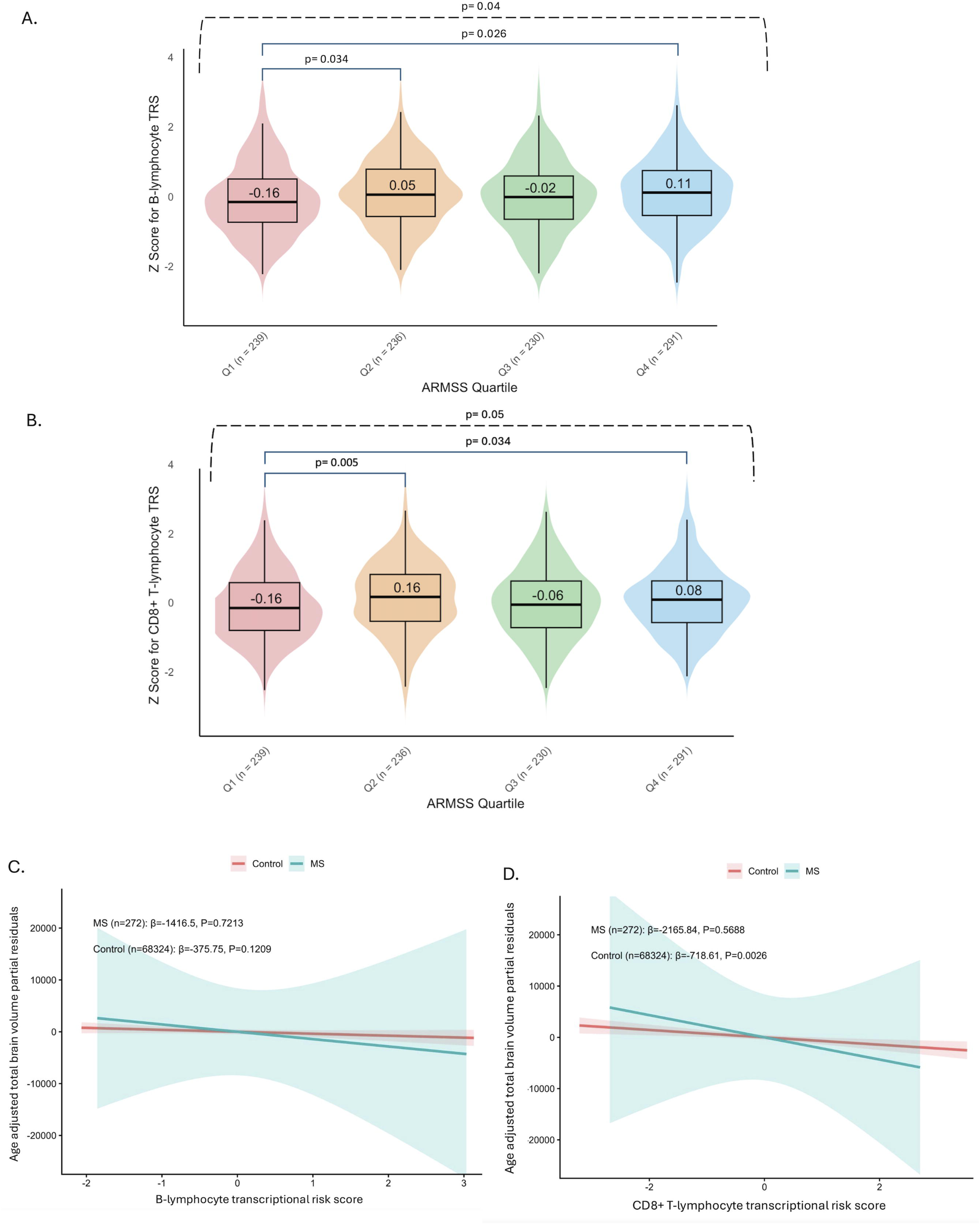
Association of standardised cell-specific transcriptional risk scores with Age-Related multiple sclerosis severity and total brain volume. **A–B:** Violin plots showing the last recorded ARMSS across quartiles of **A:** B lymphocyte and **B:** CD8+ T lymphocyte transcriptional risk scores (TRS). **C–D:** Linear regression of standardised cell specific transcriptional risk scores and age-adjusted total brain volume in UK Biobank participants, stratified by multiple sclerosis (MS) and controls **C:** B lymphocyte and **D:** CD8+ T lymphocyte transcriptional Risk scores and age-adjusted total brain volume

## Discussion

We have identified 162 novel genes putatively associated with gene expression changed linked to MS susceptibility SNP and characterized them into CNS or immune related and attributed gene expression to functional lymphocyte subsets into a transcriptional risk score. CD8+ cytotoxic lymphocyte TRS are associated with clinical MS disability (ARMSS). We also observed a similar relationship with radiological measures (smaller brain volume), as a proxy of MS severity. None of the associations were found when using conventional PRS, indicating that TRS are more sensitive to detect subgroups of pwMS.

Interestingly, the genetics of MS susceptibility has classically been considered mainly immune driven (International Multiple Sclerosis Genetics Consortium., 2019), however by integrating recent large-scale eQTL studies in CNS tissues with MS susceptibility variants, we identified several associations within brain tissue and CNS cells. Another study applying a machine learning approach demonstrated some MS susceptibility SNVs were associated with MS severity (Fuh-Ngwa et al., 2022). Moreover, we have previously shown that genomic clustering on both MS susceptibility and severity variants is able to identify subgroups of pwMS with significant differences in prognosis (Abbadessa et al., 2026, Kreft et al., 2026). This supports the view that MS susceptibility variants impact CNS tissue and long-term disease outcomes more than previously assumed.

Nevertheless, we also observed that an immune tissue TRS was associated with long-term disease outcomes. A possible explanation for this could be that ongoing chronic immune-mediated inflammation, characterised by slowly expanding MS lesions, are associated with worsening of long-term disability (Preziosa et al., 2022).

This study also highlighted that susceptibility CNS SNVs are significantly associated with both excitatory and inhibitory neurons. A recent meta-analysis of MS GWAS susceptibility variants leveraging other eQTL datasets also demonstrated increased transcriptomic expression in both excitatory and inhibitory neurons (Zeng et al., 2026). The importance of neuro-immune signalling is increasingly being recognised in MS (Woo et al., 2024). In experimental autoimmune encephalomyelitis (EAE), the animal model of MS, the interleukin (IL) 12 receptor beta on neurons has been shown to be a critical regulator of anti-inflammatory properties and prevents neurodegeneration (Andreadou et al., 2023). Interestingly, we identified *IL-12AS1* as CNS-related susceptibility gene. This anti-sense RNA regulates IL-12 production. Moreover, we also identified *IL-7* as an CNS specific MS susceptibility variant. Increased IL-7 protein expression is observed in pre-active MS lesions and CD8+ cytotoxic T-lymphocytes co-expressing the IL-7 receptor alpha chain are also found specifically in pre-active MS lesions (Kreft et al., 2012).

This study has some limitations. Although we selected from the South-Wales MS cohort only DMT naive pwMS to capture “truly” genetically-mediated severity of MS not confounded by treatment effects, this may not be fully representative for more contemporarily managed MS patients as most patients now receive DMTs during their disease course. Nevertheless, to study the potential mechanisms driving disease progression is most powerful in an untreated cohort which may more accurately reflect the underling biology of the disease and prevent challenges with statistical modelling correcting for confounding-by-indication. Despite this approach, previous studies applying TRS to Crohn’s and Alzheimer’s disease have shown greater differentiation of scores between clinically relevant subgroups (Marigorta et al., 2017; Pyun et al., 2024) than we have observed in MS. It should also be acknowledged that in the current study a significant overlap between clinical sub-groups in TRS exists. Therefore, the TRS are not as yet ready to be used in clinical practice, as is also the case with PRS (Shams et al., 2023). Thirdly, in the current study we only applied SMR to MS susceptibility GWAS, because computing PRS or TRS can be done on a single GWAS-significant hit (rs10191329) of MS progression GWAS (International Multiple Sclerosis Genetics Consortium., 2023). However, due to this, it is more likely that we underestimate rather than overestimate the number of CNS associations in MS. Finally, the UK Biobank volumetric brain imaging data was directional concordant with more advanced disease progression in the higher TRS, but these pwMS may have been treated and we were not able to adjust our models for this. Nevertheless, this may have led to an underestimation of the true effect of the transcriptional risk score (e.g. treatments reduce brain volume loss in MS and therefore partially mask the effect of genetics).

In summary, by applying a cell-type specific TRS, we have shown that higher TRS for immune cells are associated with more long-term disability in MS, most likely linked to CD8+ cytotoxic T-lymphocytes. This study showed that TRS have greater accuracy than PRS in distinguishing clinically relevant subgroups in a deeply-phenotyped real-world MS cohort and an imaging-based population study of the UK Biobank. However, this approach cannot yet be used in a clinical setting to accurately determine clinical subgroups of MS.

## Data Availability

Data from the South Wales MS Registry is available upon to eligible researchers upon signing a data transfer agreement in line with the ethical approvals. Data from the UK Biobank is available upon completion and approval of a research project and signing of a material transfer agreement. Publically available data has been cited and websites where data was obtained from have been cited in the manuscript.

## Data Availability Statement

The data used in this study is available from the corresponding author upon reasonable request, subject to signed data-sharing agreements.

## CRediT Authors contribution

**Mae Upcott:** Conceptualization, Methodology, Investigation, Formal analysis, Visualization, Writing – original draft, Writing – review and editing **Beili Shao:** Conceptualization, Methodology, Investigation, Formal analysis, Writing – original draft **Nicholas Bray**: Conceptualization, Writing-review and editing. **Sam Loveless:** Investigation, Data curation, Writing-review and editing. **Emma C. Tallantyre:** Investigation, Formal analysis, Writing – review and editing. **Neil P. Robertson:** Conceptualization, Methodology, Investigation, Formal analysis, Writing – reviewing and editing, Supervision. **Karim L. Kreft:** Conceptualization, Methodology, Investigation, Formal analysis, Writing – original draft, Writing-review and editing, Visualization, Supervision.

## Acknowledgements

We are thankful to Prof. Davey Smith, Karen Ho BSc and Dr. Tom Clark (University of Bristol, UK) for assistance with the genotyping and all people with MS who have participated in this study

## References

Abbadessa, G., Kreft, K.L., Howell, O., De Oliveira, J.V.C., Cooze, B., Papadaki, A., Jacinto, J.M., Farkas, I., Leung, Y.Y., Bonavita, S., et al., 2026. Genetic subtypes of multiple sclerosis severity are uncoupled from inflammatory lesion burden. Acta Neuropathol. 152, 24.

Andreadou, M., Ingelfinger, F., De Feo, D., Cramer, T.L.M., Tuzlak, S., Friebel, E., Schreiner, B., Eede, P., Schneeberger, S., Geesdorf, M., et al., 2023. IL-12 sensing in neurons induces neuroprotective CNS tissue adaptation and attenuates neuroinflammation in mice. Nat. Neurosci. 26, 1701–1712.

Bryois, J., Calini, D., Macnair, W., Foo, L., Urich, E., Ortmann, W., Iglesias, V.A., Selvaraj, S., Nutma, E., Marzin, M., et al., 2022. Cell-type-specific cis-eQTLs in eight human brain cell types identify novel risk genes for psychiatric and neurological disorders. Nat. Neurosci. 25, 1104–1112.

Emani, P.S., Liu, J.J., Clarke, D., Jensen, M., Warrell, J., Gupta, C., Meng, R., Lee, C.Y., Xu, S., Dursun, C., et al., 2024. Single-cell genomics and regulatory networks for 388 human brains. Science 384, eadi5199.

Fuh-Ngwa, V., Zhou, Y., Melton, P.E., van der Mei, I., Charlesworth, J.C., Lin, X., Zarghami, A., Broadley, S.A., Ponsonby, A.L., Simpson-Yap, S., et al., 2022. Ensemble machine learning identifies genetic loci associated with future worsening of disability in people with multiple sclerosis. Sci. Rep. 12, 19291.

GTEx Consortium, 2020. The GTEx Consortium atlas of genetic regulatory effects across human tissues. Science 369, 1318–1330.

International Multiple Sclerosis Genetics Consortium, 2019. Multiple sclerosis genomic map implicates peripheral immune cells and microglia in susceptibility. Science 365, eaav7188.

International Multiple Sclerosis Genetics Consortium, MultipleMS Consortium, 2023. Locus for severity implicates CNS resilience in progression of multiple sclerosis. Nature 619, 323–331.

Jacobs, B.M., Gasperi, C., Kalluri, S.R., Al-Najjar, R., McKeon, M.O., Else, J., Pukaj, A., Held, F., Sawcer, S., Ban, M., et al., 2025. Single-cell analysis of cerebrospinal fluid reveals common features of neuroinflammation. Cell Rep. Med. 6, 101733.

Jokubaitis, V.G., Campagna, M.P., Ibrahim, O., Stankovich, J., Kleinova, P., Matesanz, F., Hui, D., Eichau, S., Slee, M., Lechner-Scott, J., et al., 2023. Not all roads lead to the immune system: the genetic basis of multiple sclerosis severity. Brain 146, 2316–2331.

Kreft, K.L., Verbraak, E., Wierenga-Wolf, A.F., van Meurs, M., Oostra, B.A., Laman, J.D., Hintzen, R.Q., 2012. The IL-7Rα pathway is quantitatively and functionally altered in CD8 T cells in multiple sclerosis. J. Immunol. 188, 1874–1883

Kreft, K.L., Uzochukwu, E., Loveless, S., Willis, M., Wynford-Thomas, R., Harding, K.E., Holmans, P., Lawton, M., Tallantyre, E.C., Robertson, N.P., 2024. Relevance of multiple sclerosis severity genotype in predicting disease course: a real-world cohort. Ann. Neurol. 95, 459–470.

Kreft, K.L., Mekkes, N.J., Uzochukwu, E., Loveless, S., Wynford-Thomas, R., Harding, K.E., Wardle, M., Holmans, P., Brown, J.W.L., Lawton, M., et al., 2026. Genetic subtypes associated with multiple sclerosis severity and response to treatment. J. Neurol. Neurosurg. Psychiatry 97, 413–421.

Lappalainen, T., Sammeth, M., Friedländer, M.R., ’t Hoen, P.A., Monlong, J., Rivas, M.A., Gonzàlez-Porta, M., Kurbatova, N., Griebel, T., Ferreira, P.G., et al., 2013. Transcriptome and genome sequencing uncovers functional variation in humans. Nature 501, 506–511.

Lloyd-Jones, L.R., Holloway, A., McRae, A., Yang, J., Small, K., Zhao, J., Zeng, B., Bakshi, A., Metspalu, A., Dermitzakis, E.T., et al., 2017. The genetic architecture of gene expression in peripheral blood. Am. J. Hum. Genet. 100, 228–237.

Macnair, W., Calini, D., Agirre, E., Bryois, J., Jäkel, S., Smith, R.S., Kukanja, P., Stokar-Regenscheit, N., Ott, V., Foo, L.C., et al., 2025. snRNA-seq stratifies multiple sclerosis patients into distinct white matter glial responses. Neuron 113, 396–410.e9.

Marigorta, U.M., Denson, L.A., Hyams, J.S., Mondal, K., Prince, J., Walters, T.D., Griffiths, A., Noe, J.D., Crandall, W.V., Rosh, J.R., et al., 2017. Transcriptional risk scores link GWAS to eQTLs and predict complications in Crohn’s disease. Nat. Genet. 49, 1517–1521.

Matthews, P.M., Gupta, D., Mittal, D., Bai, W., Scalfari, A., Pollock, K.G., Sharma, V., Hill, N., 2023. The association between brain volume loss and disability in multiple sclerosis: a systematic review. Mult. Scler. Relat. Disord. 74, 104714.

Preziosa, P., Pagani, E., Meani, A., Moiola, L., Rodegher, M., Filippi, M., Rocca, M.A., 2022. Slowly expanding lesions predict 9-year multiple sclerosis disease progression. Neurol. Neuroimmunol. Neuroinflamm. 9, e1139.

Pyun, J.M., Park, Y.H., Wang, J., Bennett, D.A., Bice, P.J., Kim, J.P., Kim, S., Saykin, A.J., Nho, K., 2024. Transcriptional risk scores in Alzheimer’s disease: from pathology to cognition. Alzheimers Dement. 20, 243–252.

Qi, T., Wu, Y., Zeng, J., Zhang, F., Xue, A., Jiang, L., Zhu, Z., Kemper, K., Yengo, L., Zheng, Z., et al., 2018. Identifying gene targets for brain-related traits using transcriptomic and methylomic data from blood. Nat. Commun. 9, 2282.

Qi, T., Wu, Y., Fang, H., Zhang, F., Liu, S., Zeng, J., Yang, J., 2022. Genetic control of RNA splicing and its distinct role in complex trait variation. Nat. Genet. 54, 1355– 1363.

Scalfari, A., Neuhaus, A., Degenhardt, A., Rice, G.P., Muraro, P.A., Daumer, M., Ebers, G.C., 2010. The natural history of multiple sclerosis: a geographically based study 10: relapses and long-term disability. Brain 133, 1914–1929.

Shams, H., Shao, X., Santaniello, A., Kirkish, G., Harroud, A., Ma, Q., Isobe, N., University of California San Francisco MS-EPIC Team, Schaefer, C.A., McCauley, J.L., et al., 2023. Polygenic risk score association with multiple sclerosis susceptibility and phenotype in Europeans. Brain 146, 645–656.

Subramanian, A., Tamayo, P., Mootha, V.K., Mukherjee, S., Ebert, B.L., Gillette, M.A., Paulovich, A., Pomeroy, S.L., Golub, T.R., Lander, E.S., et al., 2005. Gene set enrichment analysis: a knowledge-based approach for interpreting genome-wide expression profiles. Proc. Natl. Acad. Sci. U. S. A. 102, 15545–15550.

Wang, X., Allen, W.E., Wright, M.A., Sylwestrak, E.L., Samusik, N., Vesuna, S., Evans, K., Liu, C., Ramakrishnan, C., Liu, J., et al., 2018. Three-dimensional intact-tissue sequencing of single-cell transcriptional states. Science 361, eaat5691.

Westra, H.J., Peters, M.J., Esko, T., Yaghootkar, H., Schurmann, C., Kettunen, J., Christiansen, M.W., Fairfax, B.P., Schramm, K., Powell, J.E., et al., 2013. Systematic identification of trans eQTLs as putative drivers of known disease associations. Nat. Genet. 45, 1238–1243.

Woo, M.S., Engler, J.B., Friese, M.A., 2024. The neuropathobiology of multiple sclerosis. Nat. Rev. Neurosci. 25, 493–513.

Yazar, S., Alquicira-Hernandez, J., Wing, K., Senabouth, A., Gordon, M.G., Andersen, S., Lu, Q., Rowson, A., Taylor, T.R.P., Clarke, L., et al., 2022. Single-cell eQTL mapping identifies cell type-specific genetic control of autoimmune disease. Science 376, eabf3041.

Zeng, L., Khan, A., Fitzgerald, K.C., Lama, T., Chen, J., Li, R., International Multiple Sclerosis Genetics Consortium, Tsai, E.A., Yu, K., Chitnis, T., et al., 2026. Genome-wide association analyses highlight the neuronal contribution to multiple sclerosis susceptibility. Nat. Genet. 58, 2177–2191.

Zheng, Z., Liu, S., Sidorenko, J., Wang, Y., Lin, T., Yengo, L., Turley, P., Ani, A., Wang, R., Nolte, I.M., et al., 2024. Leveraging functional genomic annotations and genome coverage to improve polygenic prediction of complex traits within and between ancestries. Nat. Genet. 56, 767–777.

Zhu, Z., Zhang, F., Hu, H., Bakshi, A., Robinson, M.R., Powell, J.E., Montgomery, G.W., Goddard, M.E., Wray, N.R., Visscher, P.M., et al., 2016. Integration of summary data from GWAS and eQTL studies predicts complex trait gene targets. Nat. Genet. 48, 481–487.

