## Supplementary figures for "Cell-type specific transcriptional risk scores and longitudinal multiple sclerosis outcomes"

**Fig. S1**

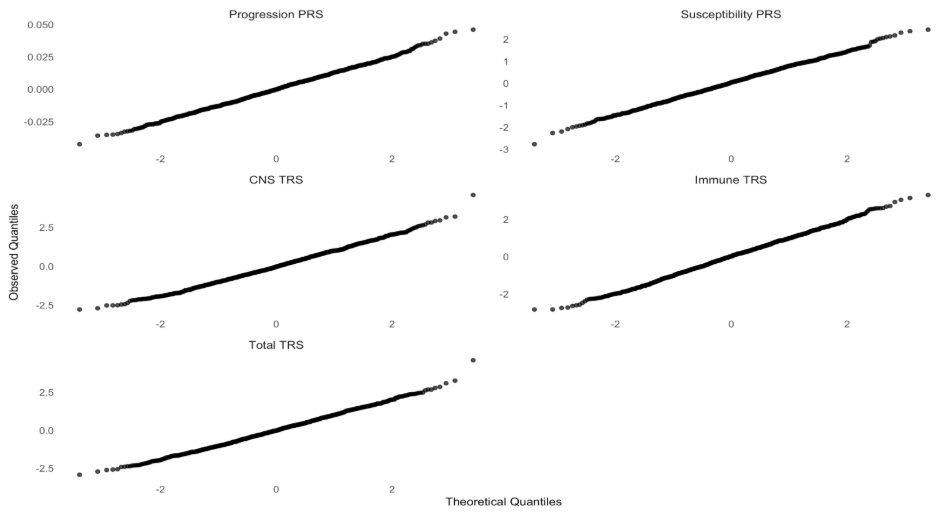

**Fig. S2**

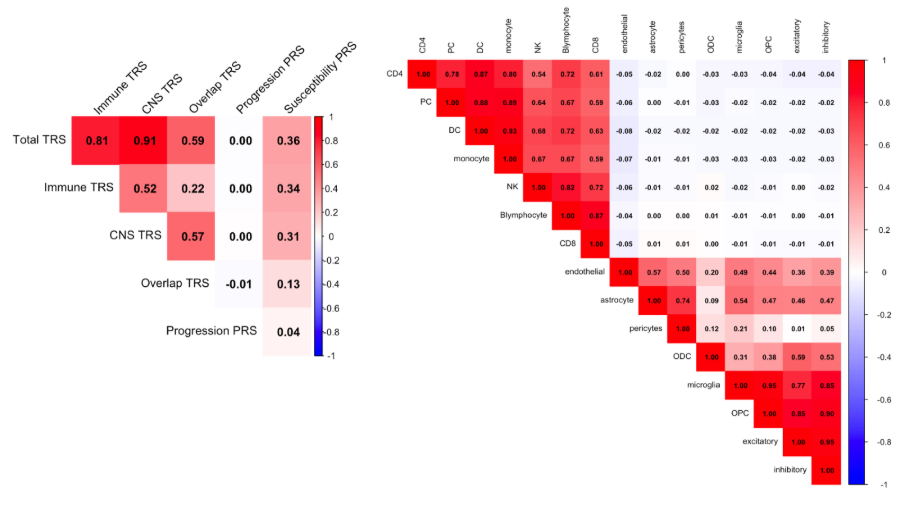

**Fig. S3**

S3A. Progression  Polygenic Risk Score

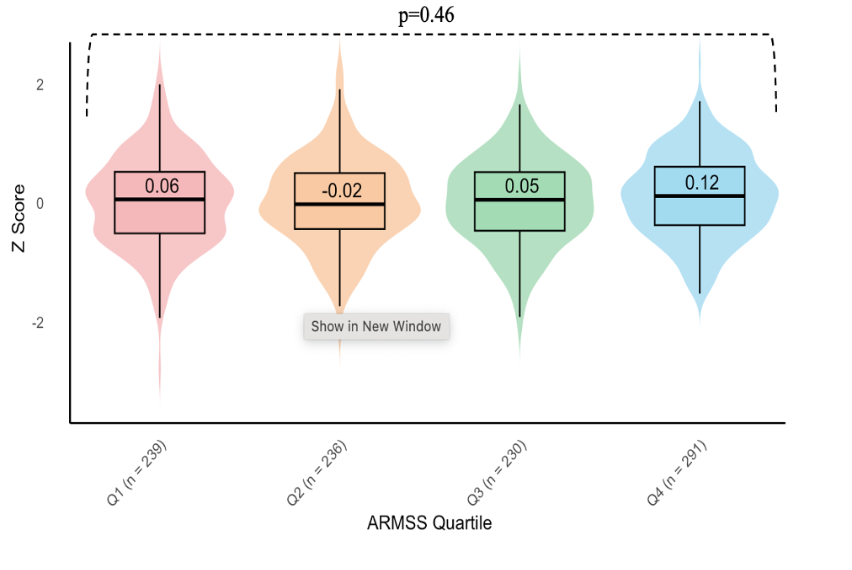

S3B. Susceptibility  Polygenic Risk Score

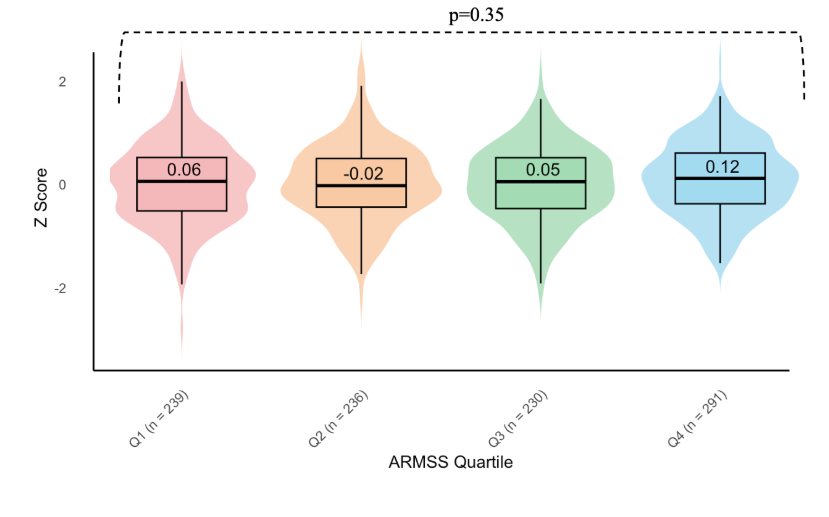

S3C. Total Transcriptional Risk score

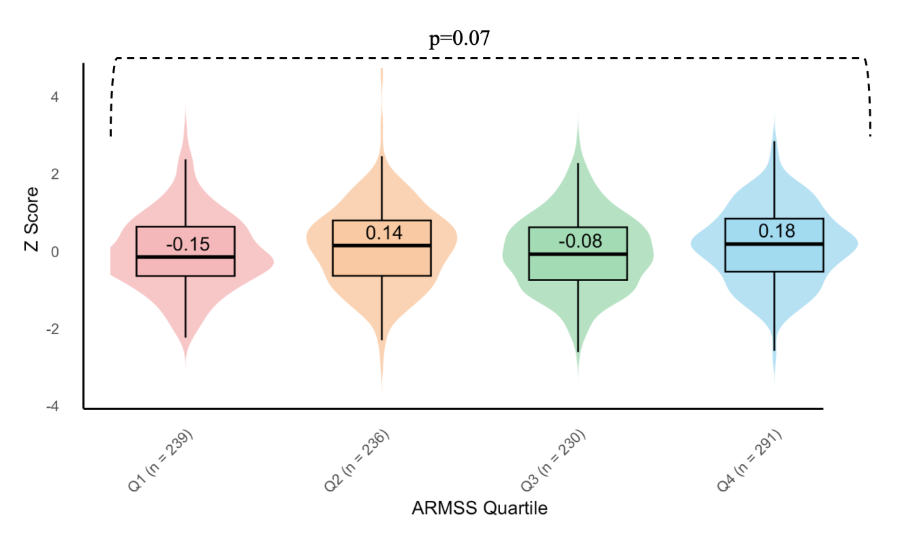
 S3D. CNS Total transcriptional risk score

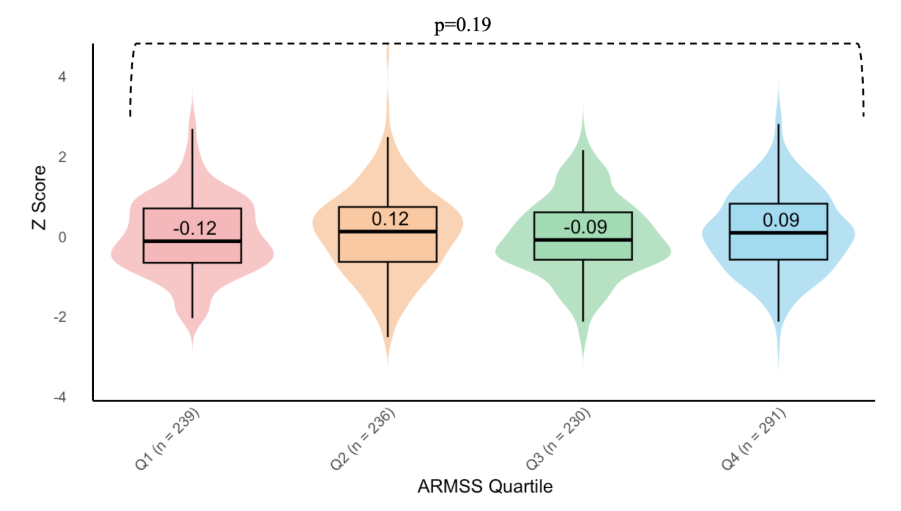

**Fig. S4**

S4a. Susceptibility Polygenic Risk Score

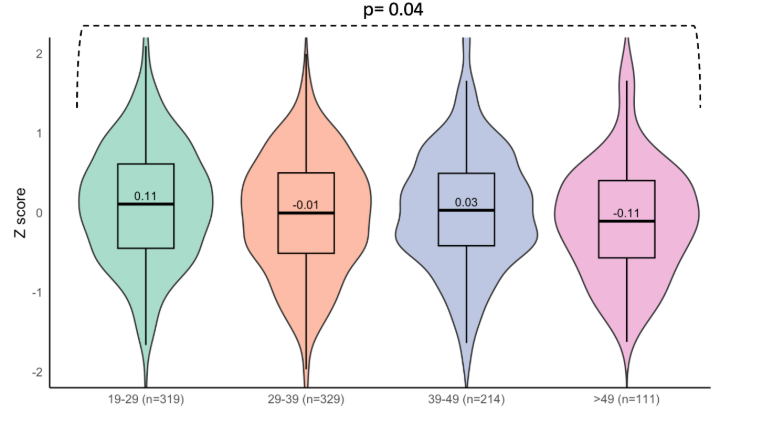

S4b. Progression Polygenic Risk Score

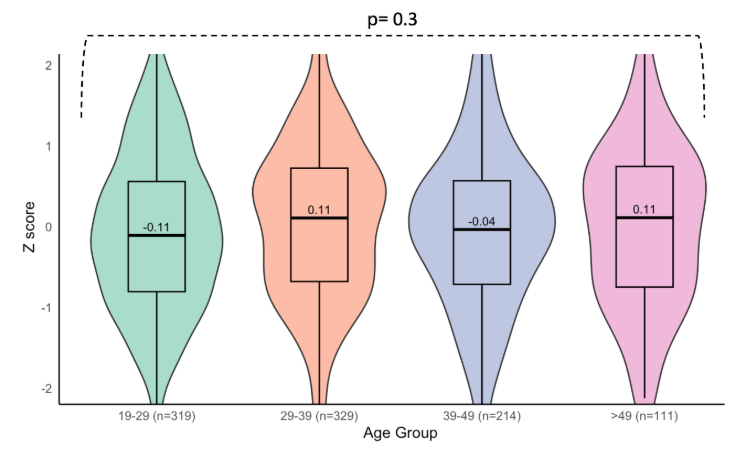

S4c. Total transcriptional risk score

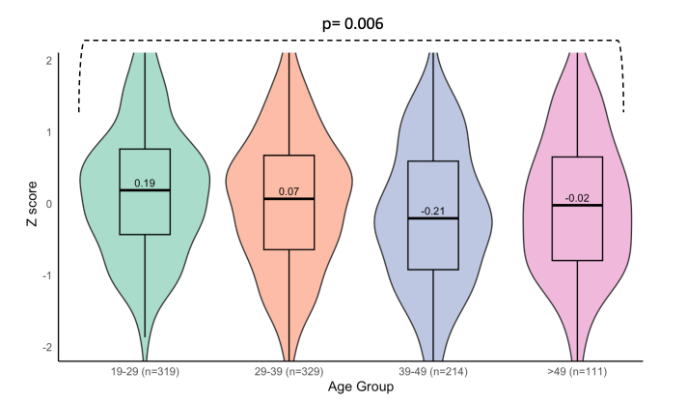

S4d. Immune transcriptional risk score

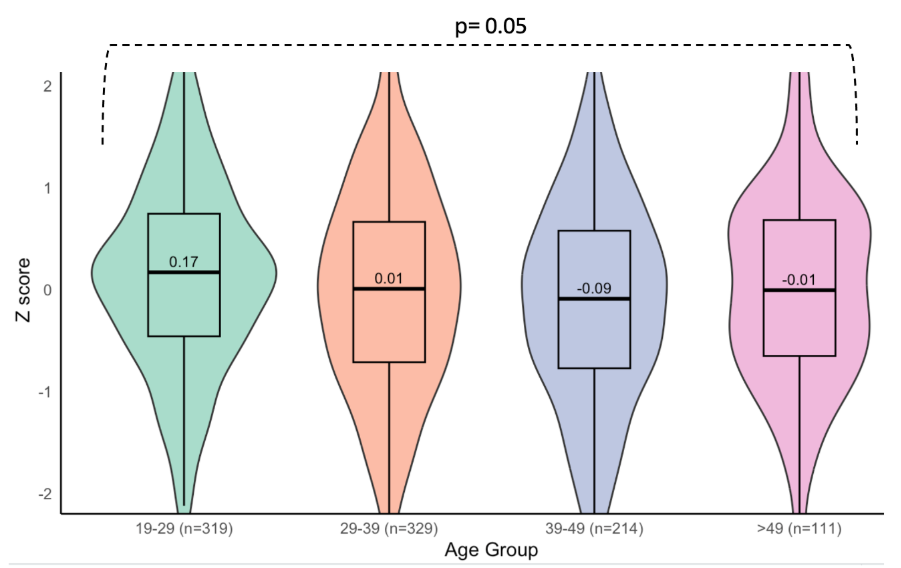

S4e. CNS transcriptional risk score

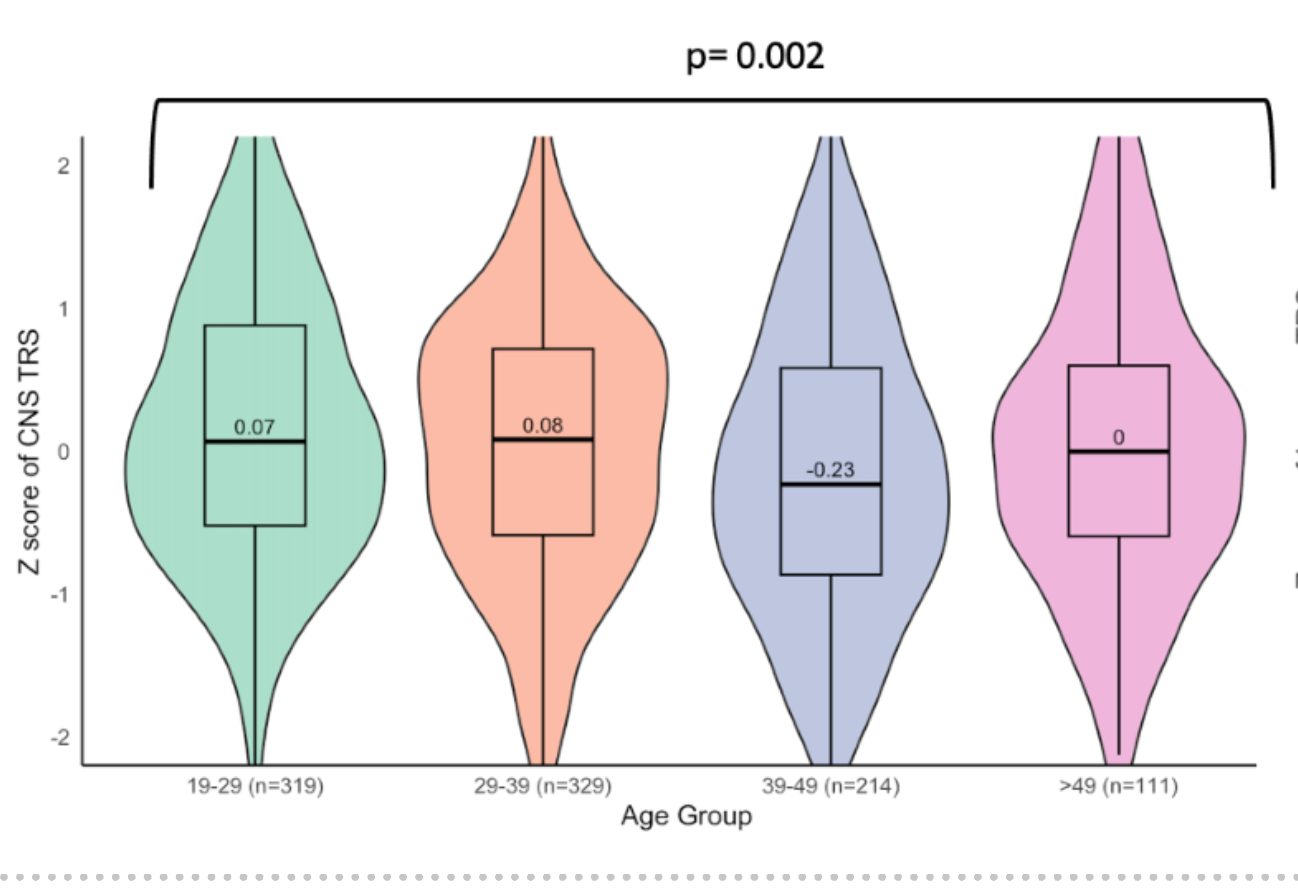
