## Supplementary table 1 and 2 for "Cell-type specific transcriptional risk scores and longitudinal multiple sclerosis outcomes"

Table S1. Target SNVs which could not be identified within in the South Wales MS Registry genotype data.

| SNP | Chromosome | Position |
| --- | --- | --- |
| rs2298210 | 1 | 1211473 |
| rs12097268 | 1 | 2545919 |
| rs58055515 | 1 | 2691022 |
| rs617599 | 1 | 160193169 |
| rs3747623 | 1 | 160193719 |
| rs2984920 | 1 | 192575665 |
| rs4254532 | 2 | 191114452 |
| rs13197301 | 6 | 135497787 |
| rs7833924 | 8 | 143921861 |
| rs7925844 | 11 | 302505 |
| rs34481144 | 11 | 320836 |
| rs7944394 | 11 | 323649 |
| rs6598042 | 11 | 324635 |
| rs11246067 | 11 | 326564 |
| rs7396777 | 11 | 328652 |
| rs6598039 | 11 | 330973 |
| rs34702437 | 11 | 332798 |
| rs4758639 | 11 | 305406 |
| rs61876251 | 11 | 329896 |
| rs7942247 | 11 | 316349 |
| rs1059501 | 12 | 6451407 |
| rs12296430 | 12 | 6394334 |
| rs8009897 | 14 | 100358226 |
| rs9704354 | 14 | 20758103 |
| rs2074585 | 15 | 90466252 |
| rs8059619 | 16 | 29914124 |
| rs4407079 | 16 | 29920578 |
| rs7184288 | 16 | 29923731 |
| rs4424923 | 16 | 29927972 |
| rs4527039 | 16 | 29935624 |
| rs3814883 | 16 | 29983601 |
| rs28529403 | 16 | 30123335 |
| rs620238 | 16 | 1003817 |
| rs11647753 | 16 | 29916794 |
| rs12716973 | 16 | 29926331 |
| rs2289292 | 16 | 30086309 |
| rs6565175 | 16 | 30133058 |
| rs3809624 | 16 | 30091481 |
| rs55732507 | 16 | 30130664 |
| rs11669861 | 19 | 19166593 |
| rs461709 | 19 | 47163454 |
| rs884171 | 19 | 47213090 |
| rs10424211 | 19 | 865818 |
| rs1683593 | 19 | 867735 |
| rs2072563 | 19 | 46023390 |
| rs2298432 | 22 | 21768900 |
| rs4821544 | 22 | 36862461 |

Table S2. Proxy SNVs in perfect linkage disequilibrium used in South Wales MS Registry genotype data

| SNP | Chromosome | Position | Proxy SNP |
| --- | --- | --- | --- |
| rs7155 | 8 | 143915294 | rs7014582 |
| rs1570305 | 14 | 100341818 | rs59768891 |
| rs941927 | 14 | 100374476 | rs58659361 |
| rs6889220 | 5 | 177266887 | rs56500479 |
| rs11993233 | 8 | 143928115 | rs62521874 |
